# Dietary Factors and Predominant Eye Diseases in Sub-Saharan African Populations: A Comprehensive Systematic Review Protocol

**DOI:** 10.1101/2025.02.17.25322272

**Authors:** Isaiah Osei Duah, Kwadwo Owusu Akuffo, Josephine Ampong, David Owiredu, Anthony Danso-Appiah

## Abstract

**Background:** Evidence linking diet and ocular diseases is growing, yet variations persist, with a paucity of data in sub-Saharan Africa. The proposed review will systematically synthesize evidence on dietary factors associated with predominant eye disorders (cataracts, refractive error, glaucoma, diabetic retinopathy, age-related macular degeneration, and dry eye disease) in the sub-Saharan African population.

**Methods:** The systematic review and meta-analysis will follow the gold standard for reporting systematic reviews and meta-analysis, thus Preferred Reporting Items for Reporting Systematic Reviews and Meta-analysis (PRISMA), and Meta-analysis of Observational Studies in Epidemiology (MOOSE) guidelines. The study will identify all relevant studies published, unpublished, and pre-prints from PubMed, SciVerse Scopus, Web of Science, Google Scholar, Embase, Health Inter-Network Access to Research Initiative (HINARI), Cumulative Index to Nursing and Allied Health Literature (CINAHL), African Journal of Science, Biomed Central, and Cochrane Library and relevant institutional databases without date restrictions. We will check the reference lists of all studies identified by the methods and contact experts for additional relevant studies. Data will be analyzed with the Review Manager (RevMan) software. The random effects model will be used to pool study effect estimates and robustness assessed through sensitivity analyses. Heterogeneity across studies will be assessed using the I^2^-statistic. Publication bias will be investigated graphically with a funnel plot and statistically with Egger’s regression intercept method or Begg’s rank correlation tests. All estimates will be reported with their confidence intervals, and statistical analysis will be set at a significance level of p<0.05. The results of the review will be presented based on the PRISMA guidelines.

## Background

The burden of eye diseases and complications of blindness has increased exponentially and remains a public health threat globally [1], although it differs across geographical regions. Uncorrected refractive errors, cataracts, glaucoma, diabetic retinopathy, age-related macular degeneration, and dry eye diseases remain consistently high and account for millions of moderate-to-severe vision impairments and blindness [2–5]. Most cases occur in low-and-middle-income countries (LMICs) and territories [6, 7]. In sub-Saharan Africa, for example, four million people live with blindness, and about 18 million have moderate-to-severe vision impairment [4]. The trends of ocular diseases within the region reflect a growing public health concern that necessitates urgent action.

In particular, cataract, characterized by cloudiness or opacification of the lens; uncorrected refractive errors, defined as an imbalance between axial length and refractive power of the eye; glaucoma, explained by progressive optic neuropathy with associated visual field changes; diabetic retinopathy, delineates microvascular end-organ complication of diabetes mellitus; age-related macular degeneration expounds the deterioration of the macula and/or fovea; dry eye diseases describes as instability in tear film production, represents a major health, economic, and societal challenges within this population[4, 8–10]. For instance, in children, vision impairment from uncorrected refractive error has an adverse effect on their academic prowess, self-image, and social relations[11–14]. Similarly, vision loss affects young adults and/or middle-aged individuals’ career aspirations, employment prospects, and socioeconomic status[15]. Likewise, in the older or aged population, derangement in vision culminates in physical dependency and psychological distress[16, 17]. Most probably, a deficit in vision function reversibly affects the overall quality of life across the entire age spectrum.

Several factors, notably genetics[18–20], environment[21–23], and lifestyle changes[24–26], engage in complex interplay in the pathogenesis of eye diseases. On the one hand, nutrition and diet are commonly implicated as modifiable risk factors [27]. Dietary factors, including macronutrients and micronutrients, are critical in organ system development, nourishment, and function. The food nutrients possess biologically potentiation features such as antioxidant[28, 29], anti-inflammatory[29], anti-angiogenic[29], anticarcinogenic[30, 31], neuroprotection[29] and light filtering properties[32, 33], which putatively protect against debilitating ocular diseases. In addition, they terminate free radical reactions ensuing oxidative stress, and lipid peroxidation hypothesize to mediate most ocular diseases[34]. Given the underlying central dogma, we postulate that dietary factors could serve as an impetus to cause a reduction in the prevalence and incidence of ocular diseases in sub-Saharan Africa when evidence about the protective quantities is made available.

Dietary factors are fundamental in human nutrition. However, controversies remain about its adequacy [35, 36], which limits its direct applicability in ocular health and vision medicine[37–40]. Evidence from various systematic reviews and longitudinal studies is diverse[39–47]. Thus, whereas some authors report an association between higher intake and reduced risk of specific ocular diseases[42, 43], others report otherwise[39, 41, 44, 48], with most reviews showing no significant association[46–49], and a fewer more systematic reviews unable to draw a robust conclusion[29, 38, 40, 45, 46]. Despite the comprehensive nature of the above reviews, which provide valuable insights, its focus was not on the sub-Saharan population, which conceals its directedness. Dietary diversity recognized as proxies for micronutrient sufficiency, quality, and adequacy of diets[50] are linked to socioeconomic status[51]. The variations in the standard of living in sub-Saharan Africa compared with Northern and Southern Africa terrains and the westernized world imply differences in health status variables and dietary indicators. Corroborated by a multi-center cross-sectional study, Galbete et al. revealed dietary differences among sub-Saharan migrants and their local compatriots. Whiles the latter group showed diverse eating patterns, the former category mainly consumed starchy staples and diets dominated by animal or animal-based products[52]. The acculturation, urbanization, and globalization consistently compound the dietary habits of the sub-regional inhabitants, which implies that ascendancies in the burden of the above ocular pathologies are expected if any concrete interventional strategies are implemented.

Nonetheless, until substantial progress is made to curtail the regional crisis a systematic synthesis of evidence is presently warranted. The robustness, transparency, minimally bias, and the inherent methodical rigor of this approach will bring to light the unknown, summarize the known, show masked trends, distilled insights and provides a simple comprehensive regionally based recommendation for clinicians and policymakers. Further, the outcome of the study will transform nutritional policies geared towards reducing the burden of eye diseases in the region. The overarching goal of the study is to systematically review the literature and synthesize evidence on the dietary factors linked with the predominant ocular diseases in sub-Saharan Africa. Specifically, the review aims to answer the question, “What dietary factors are associated with predominant eye diseases (cataracts, refractive errors, glaucoma, diabetic retinopathy, age-related macular degeneration, dry eye disease) in the sub-Saharan African population?”

## Methods

The prepared protocol is consistent with the Meta-analysis for Observational Studies in Epidemiology (MOOSE)[53] and Preferred Reporting Items for Systematic Reviews and Meta-Analyses Protocol (PRISMA-P) guidelines [54, 55], and the findings from the study will be presented following the Preferred Reporting Items for Systematic Reviews (PRISMA) guidelines [56, 57]. The protocol was registered in the International Prospective Register for Systematic Reviews (ID: CRD42023402042) before the commencement of the full systematic review.

## Criteria for Considering Studies for Inclusion or Exclusion in the Review

The criteria for inclusion follow PICOS elements: population, intervention, comparators, outcomes, and study design.

## Population

The study population will be all human participants of all ages living in sub-Saharan Africa that have been clinically diagnosed or self-reported to have either one or both eyes affected by either cataract, refractive errors, glaucoma, diabetic retinopathy, aged-related macular degeneration, dry eye disease.

## Intervention

Interventions of the selected studies will examine the effect and/or association of dietary factors (saturated fatty acids, unsaturated fatty acids, unsaturated-fat-diet, monosaturated fatty acids, polysaturated fatty acids, omega-3-fatty acids, omega-6-fatty acids, omega-9-fatty acids, oleic acids, linoleic acids, alpha-linoleic acids, modified-fats, low-fats, trans fat, cholesterol, carbohydrates, refined carbohydrates, calories, glycemic index, caloric restrictions, energy-restricted diets, sugar, proteins, dietary proteins, vitamins, vitamin a, vitamin K, vitamin D, vitamin E, vitamin B 1, vitamin B 2, vitamin B 3, vitamin B 6, vitamin B 12, vitamin H, carotene, thiamine, riboflavin, niacin, biotin, pantothenic acids, pyridoxine, folate, cobalamine, minerals, sodium, potassium, magnesium, iodine, selenium, zinc, copper, iron, special nutrients tea, coffee, caffeine, milk, fibre, oily fish, fruits, fruit juice, vegetables, salt, nuts, antioxidants, dark green, leafy vegetables, green leafy vegetables, meat, bread, cereals, lutein, zeaxanthin, carotenoids, alpha-carotene, β-carotene, β-cryptoxanthin, alpha-tocopherol, curcumin, saffron, resveratrol, lycopene, glutathione, nitric oxide, flavonoids, ginkgo biloba extract, plant-based diet, alcohol, skim milk, poultry, non-meat animal products, Red Meat, Vegetable Oils, Animal Fats, Spinach, Collard Greens) to decrease either the prevalence, incidence, severity, and progression of the predominant eye diseases (cataract, uncorrected refractive errors, glaucoma, diabetic retinopathy, age-related macular degeneration, and dry eye disease).

## Comparators

Comparators for the study where appropriate will be control arm that takes the recommended dietary intake, dietary supplement, pharmacological and non-pharmacological interventions to mitigate the abovementioned eye diseases.

## Outcomes

The primary outcomes will be reduction of prevalence, incidence, severity and/or progression of the predominant eye diseases (cataract, uncorrected refractive errors, glaucoma, diabetic retinopathy, age-related macular degeneration, and dry eye disease). The study will evaluate secondary outcomes that comprises contrast sensitivity, visual acuity, pin hole acuity, glare, macular pigment optical density, color vision, night blindness, retinitis pigmentosa, visual acuity, pinhole acuity.

## Adverse Events

- Non serious adverse events
- Serious adverse events (example vision impairment, decreased photopic and scotopic vision, retinopathy, retina degeneration, optic neuropathy) evaluated using standardized procedures or diagnostic equipment.

## Study design

Eligible studies should present empirical findings on linkage between dietary factors (other than supplement) and ocular diseases among human subjects in sub-Saharan Africa. Specifically, studies to be included should have the following designs (1) randomized control trials [RCTs], (ii) prospective cohort study, (iii) case control studies, (iv) cross-sectional studies. Data from national health examination surveys without clearly defined denominator, systematic reviews, opinions, commentaries will be excluded. Qualitative studies, case studies, case series will be included for exploratory purposes and in-depth analysis on the interaction between the intervention and outcomes, however, they will not be included for meta-analysis. Studies should give the zero-order relationships between dietary factors and ocular diseases or provide adequate information for these associations (effect sizes) to be estimated in order to be included in the meta-analysis.

## Inclusion criteria

The systematic review will include studies based on the study design, study population, interventions, comparators, and outcomes described above.

## Exclusion criteria

The systematic review will exclude studies that utilize data from national health examination surveys, systematic reviews, meta-analysis, overviews of reviews, disproportionate or heterogeneous distributions. Furthermore, studies that comprise non-human subjects, conducted outside sub-Saharan Africa, and in primary literature in languages other than English will be excluded.

## Data Sources, Search Terms, and Search Strategies for Identifying Studies

A comprehensive search strategy will identify relevant studies (published, unpublished and pre-prints) from Public/Publisher PubMed, SciVerse Scopus, Web of Science, Google Scholar, Health Inter-Network Access to Research Initiative (HINARI), Cumulative Index to Nursing and Allied Health Literature (CINAHL), African Journal of Science, African Journals Online, BioMed Central, Cochrane Library and relevant Institutional databases. There will not be language and date restrictions. Medical subject headings (MeSH) and word list will be used in conjunction with appropriate Boolean operators; OR” and “AND” (as exemplified in Table 1 for PubMed database) and adjusted appropriately for the remaining database.

**Table 1:**
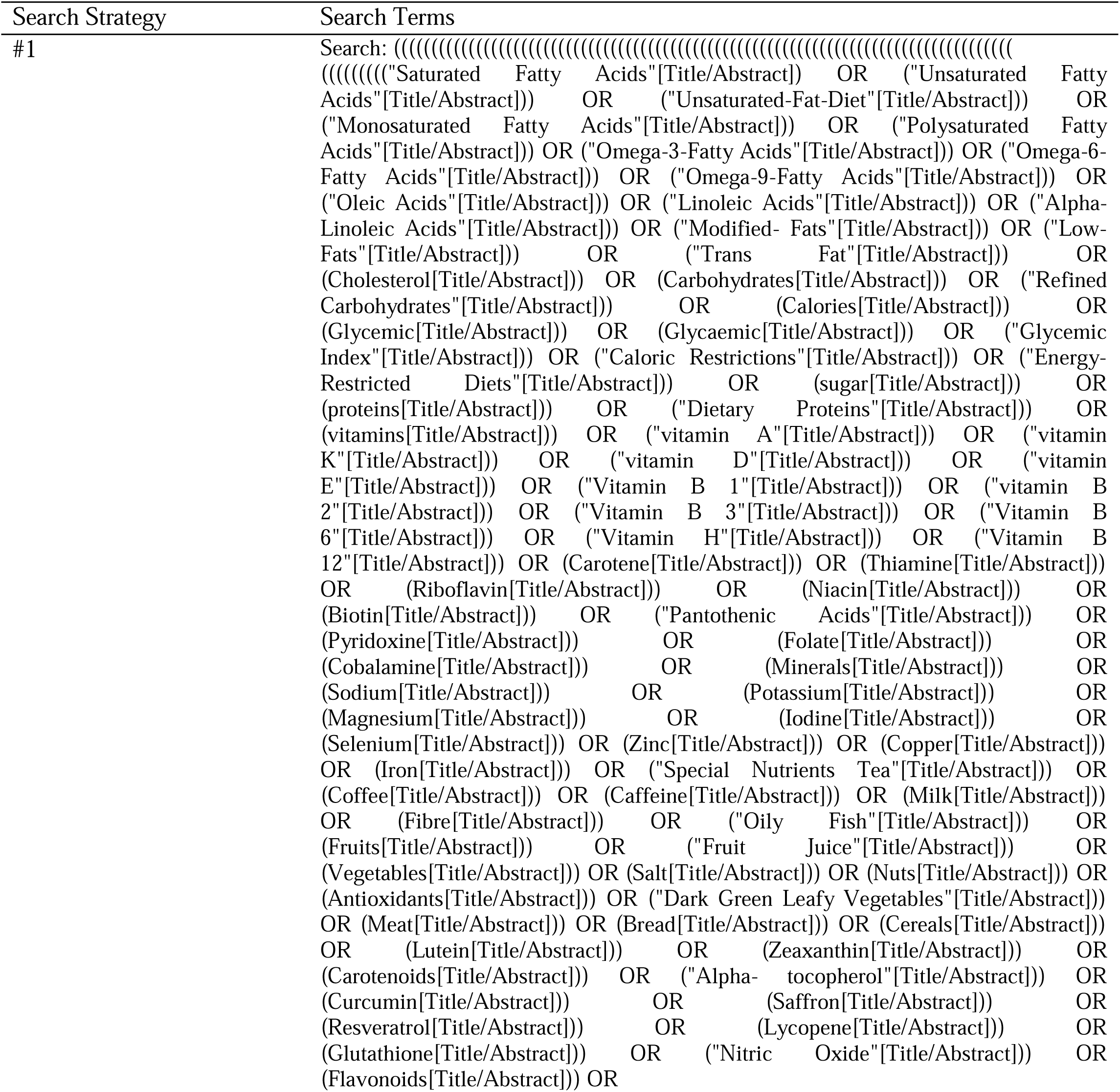
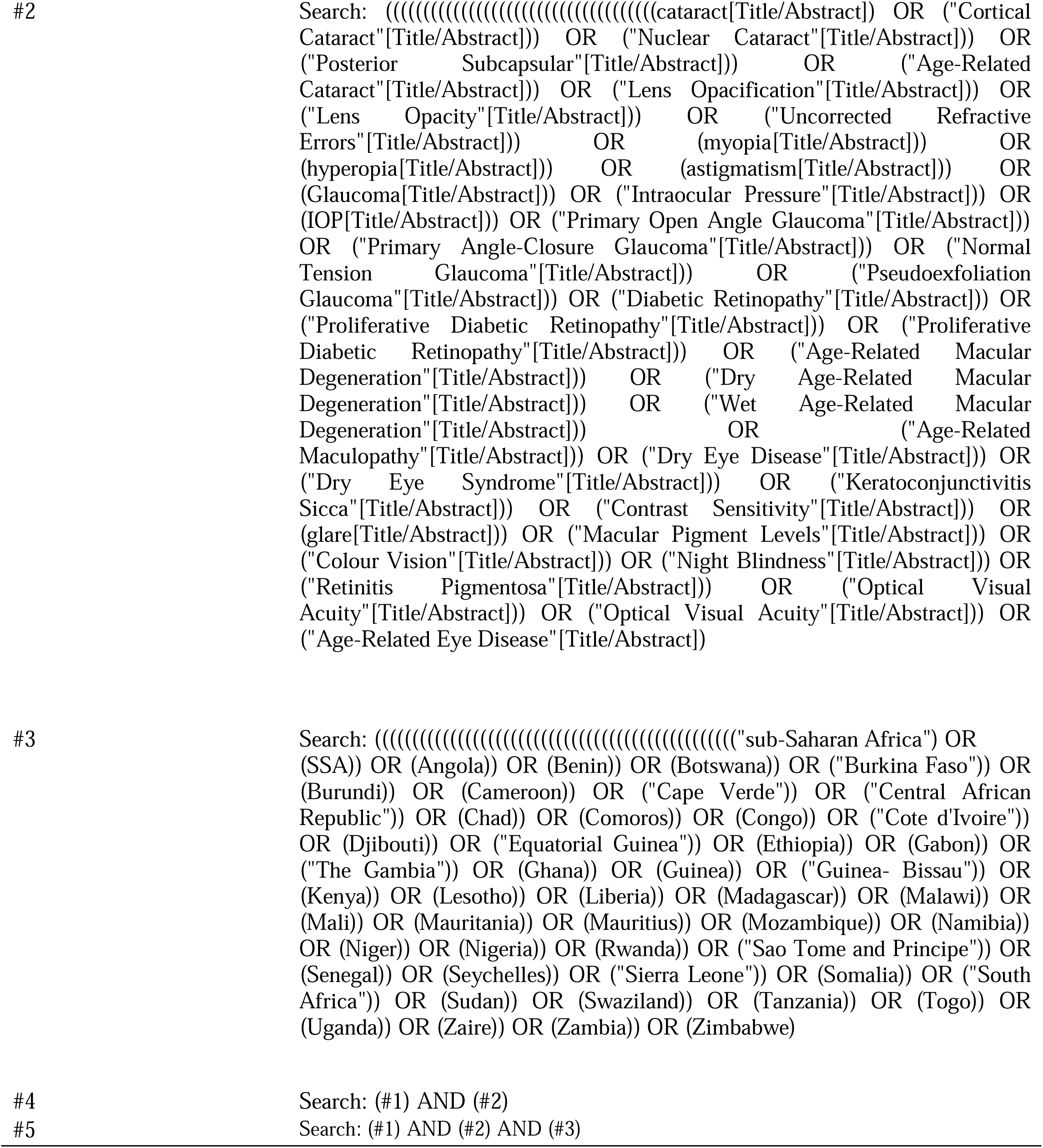
Search Strategy for PubMed.

## Screening and Selection of Studies

The retrieved articles and abstract will be imported into Endnote version 9 and references manage. Duplicates will be removed entirely and subsequently de-duplicated files imported into Rayyan Intelligent Systematic Review toolbox [58] for screening. A hierarchical three-dimensional approach will be employed in the screening process. First, articles will be assessed based on titles and abstracts, secondly, the full text-version will be critically evaluated for methodological quality and presence of effect sizes. Concurrently, following a prespecified eligibility criteria, authors will include all feasible studies. Of note to eliminate bias and enhance transparency all included papers will be separately and independently assessed all the authors. Where there is indecisiveness and/or where the lead student researcher included a study but one or more of the experts excluded, a consensus will be reached on that study. To agree on a decision a vote will be cast for inclusion or exclusion and in a 3:1 ratio in favor of a ‘yes’ or a ‘no’. On the other hand, if there is an equal proportion in votes on a decision, researchers will seek the opinion of an independent systematic reviewer before that study is either included or excluded. All methodological approach and decisions that are taken prior to the selection of our analytical sample will be documented in a PRISMA flow diagram.

## Data Extraction and Management

The review will be conducted using Review Manager version 5.4. At least two reviewers will extract data using pre-tested data extraction tool. The data to be extracted will include study characteristics such as authors name, year of publication, title, country, primary focus of the review question i.e., dietary factors and ocular diseases, review questions in terms of sample size, population, intervention, comparator, outcomes, and study design. Dichotomous outcomes: odds ratios and/or relative risk as well as their 95% confidence interval values will be extracted whereas mean differences and standard deviations will be obtained for continuous variables. All data will be consolidated in an excel spreadsheet. Any disagreements will be resolved through discussion.

## Quality Assessment

Randomized controlled trials (RCTs) bias will be used to assess using the risk of bias in each trial based on the Cochrane Risk of Bias tool. Six domains will be assessed including sequence generation, allocation concealment, blinding (of participants, personnel, and outcome assessors), incomplete outcome data, selective outcome reporting; and other sources of bias. We will categorize these judgments as low, high, or unclear risk of bias. Risk of bias in non-randomized studies of Interventions (ROBINS-I) tool will be to assess the risk of bias in observation quantitative studies. Seven domains of bias will be assessed are confounding bias, participant selection bias, interventions classification bias, deviations from intended interventions bias, missing data bias, measurement of outcomes bias and selection of the reported result bias. We will rate the risk of bias as low, moderate, serious, or critical. Confidence in the Evidence from Reviews of Qualitative research (GRADE CERQual) approach will be used to assess the confidence in each study. The domains that will be assessed are studies’ methodological limitations, coherence of the review study, data contributing adequacy, and included studies’ relevance. We will judge the confidence as high, moderate, low, or very low.

## Data Analysis

Concurrently, comparisons between prospective datasets and data from cross-sectional studies will be investigated using moderation analyses. Forest plots will be generated for the various comparisons. The effect sizes across the various studies will be pooled using the random effects model. The robustness of the pooled estimates and the magnitudes of the individual studies will be examined using sensitivity analysis. Heterogeneity within the studies will be assess using the Cochrane Q_within_ – statistics and χ² at a significance level of 0.05 and percentage heterogeneity will be evaluated using the Higgins I² index. Similarly, any anticipated publication bias will be assessed graphically using funnel plot and statistically using Egger’s regression intercept method and/or Begg’s rank correlation tests. Given the variations in the age categories of our sample a sub-group analysis will be investigated using meta-regression analyses.

## Dealing with Missing Data

We planned to perform analysis only completed data, however, incomplete datasets will be excluded.

## Measures of Effect

Dichotomous outcomes in the study will be evaluated using risk ratios and/or odds ratio whereas mean differences (MD) and geometric mean ratios will serve as measures of effects to condense continuous outcomes respectively summarized in arithmetic or geometric means. All statistical analysis will be performed at a significance of p < 0.05.

## Assessment of Heterogeneity

Comprehensive sub-group analysis will be performed to address significant potential heterogeneity (I²> 50% and/or p < 0.10) that may arise from differences in eye conditions; age categories (children, adults, and aged); diagnoses (self-reported, clinician-assessed), study designs. Heterogeneity will be assessed using the I² statistic and Cochran’s Q test. If significant heterogeneity is detected (I²> 50% and/or p < 0.10), subgroup analyses will be conducted to explore the potential sources of heterogeneity.

## Sensitivity Analyses

Sensitivity analyses will be conducted to assess the impact of individual studies on the overall effect size and to evaluate the robustness of the results.

## Meta-Regression

Meta-regression analyses will be conducted to investigate potential sources of heterogeneity.

## Patient and Public involvement

Patients or the public were not involved in the design, or conduct, or reporting, or dissemination plans of our research.

## Discussion

The aim of the planned review is to systematically synthesize evidence on dietary factors associated with ocular diseases in sub-Saharan Africa. Diet and dietary metabolites constitute the building block of most living cell and the foundational elements for several biological reactions within the body including the famous Emden Meyerhof/ glycolytic pathway, Krebs/tricarboxylic cycle, electron transport chain and the one-carbon metabolic cycle. They serve as cofactors and starters of varying reaction machineries which reversely prevent oxidative stress a mechanisms hypothesized to mediate the pathogenesis of cataract, refractive errors, glaucoma, diabetic retinopathy and glaucoma. As a results of their free radical scavenging properties, inherent ability to terminate lipid peroxidation and subsequently restore membrane structures and function they are hypothesized to confer protective effects on varying on eye diseases including those highlighted in our review. Nevertheless, various systematic reviews conducted in various population are inconclusive and there is paucity of data in sub-Saharan Africa. Hence the outcome of the data will provide regional evidence, and recommendations from the review will inform ophthalmic clinicians and general physicians on the various dietary factors with beneficial and deleterious effects on the eyes which will invaluably help them advise their clients. Further, the results will guide policy makers to develop interventions to reduce the burden of eye disease in this part of the world. This review has several strengths and some limitations worth highlighting. For instance, this will be the first review to systematically synthesize evidence on the dietary factors that influence predominant eye diseases within the sub-Saharan African population. Second, the study will employ a more robust strategy that follows the PRISMA an/or MOOSE guidelines where appropriate. Contrary, despite using multiple databases there remain a possibility of missing out on studies not indexed in these databases. However, this indexing bias is underscored by the review following a prespecified inclusion and exclusion criteria together with no date and language restrictions that extends the possibility of including all potential studies.

## Data Availability

No datasets were generated or analysed during the current study. All relevant data from this study will be made available upon study completion.

## Acknowledgement

This systematic review protocol has been prepared as part of capacity building initiative by the Centre for Evidence Synthesis and Policy (CESP), University of Ghana and Africa Communities of Evidence Synthesis and Translation (ACEST) that train experts in evidence synthesis and translation across low income and middle-income countries (LMICs), particularly Africa. The lead author, Isaiah Osei Duah Junior, received mentorship from the Senior author, Professor Anthony Danso-Appiah; the Director, Centre for Evidence Synthesis and Policy, University of Ghana, Accra).

## Ethics and dissemination

This systematic review and meta-analysis will rely on secondary data available in scientific databases, hence ethical clearance is not applicable. The findings of this study will be disseminated through a peer-reviewed journal and scientific conferences.

## Authors contribution

IODJ, ADA, and KOA conceptualized the study; IODJ, KOA, JA, DO, and TDA contributed to the methodology section; IODJ, DO, and ADA develop the search strategy; IODJ, ADA, JA, and KOA drafted the original protocol; IODJ, ADA, JA, DO, and KOA critically reviewed for important intellectual merits; IODJ, ADA, and KOA administered the project; TDA supervised the study. All authors approved the publication of the protocol.

## Competing interests

The authors declare no competing interests.

**S1 Table:** PRISMA-P (Preferred Reporting Items for Systematic Review and Meta-Analysis Protocols) 2015 checklist: Recommended items to address in a systematic review protocol

